# Whole-exome imputation within UK Biobank powers rare coding variant association and fine-mapping analyses

**DOI:** 10.1101/2020.08.28.20180414

**Authors:** Alison R. Barton, Maxwell A. Sherman, Ronen E. Mukamel, Po-Ru Loh

## Abstract

Exome association studies to date have generally been underpowered to systematically evaluate the phenotypic impact of very rare coding variants. We leveraged extensive haplotype sharing between 49,960 exome-sequenced UK Biobank participants and the remainder of the cohort (total *N*~500K) to impute exome-wide variants at high accuracy (*R*^2^>0.5) down to minor allele frequency (MAF) ~0.00005. Association and fine-mapping analyses of 54 quantitative traits identified 1,189 significant associations (*P*<5 x 10^-8^) involving 675 distinct rare protein-altering variants (MAF<0.01) that passed stringent filters for likely causality; 600 of the 675 variants (89%) were not present in the NHGRI-EBI GWAS Catalog. We replicated the effect directions of 28 of 28 height-associated variants genotyped in previous exome array studies, including missense variants in newly-associated collagen genes *COL16A1* and *COL11A2*. Across all traits, 49% of associations (578/1,189) occurred in genes with two or more hits; follow-up analyses of these genes identified long allelic series containing up to 45 distinct likely-causal variants within the same gene (on average exhibiting 93%-concordant effect directions). In particular, 24 rare coding variants in *IFRD2* independently associated with reticulocyte indices, suggesting an important role of *IFRD2* in red blood cell development, and 11 rare coding variants in *NPR2* (a gene previously implicated in Mendelian skeletal disorders) exhibited intermediate-to-strong effects on height (0.18-1.09 s.d.). Our results demonstrate the utility of within-cohort imputation in population-scale GWAS cohorts, provide a catalog of likely-causal, large-effect coding variant associations, and foreshadow the insights that will be revealed as genetic biobank studies continue to grow.

## INTRODUCTION

Exome association studies have observed that rare coding variants tend to have larger phenotypic effects than common variants and collectively contribute an important component of complex trait heritability^1–4^. However, the phenotypic effects of very rare coding variants have been difficult to comprehensively assess, as exome sequencing studies have not yet reached the sample sizes needed to power such analyses (*N*>100,000)^5-10^, and imputation of rare variants into cohorts of this scale has been insufficiently accurate^11^. The largest exome-wide association studies conducted to date have analyzed cohorts of *N*~50,000 exome-sequenced individuals, and while these studies have identified modest numbers of variants and genes associated with phenotypes, they have largely been underpowered to evaluate the effects of individual very rare coding variants^7–10^.

The UK Biobank (UKB) is a powerful resource for genetic association analyses because of its large sample size (N~500,000) and deep phenotyping^12^. Previous studies of UKB have examined disease associations of protein-truncating variants genotyped on the UK Biobank array, which was designed to include the majority of predicted loss-of-function (LoF) variants with MAF>0.02% and missense variants with MAF>0.2%^13,14^. However, most LoF variants are ultra-rare (MAF<0.01%), such that only ~14% of rare LoF variants detected in whole-exome sequencing (WES) of 49,960 UKB participants had been genotyped on the UK Biobank array^8^.

We reasoned that although exome sequencing of ~10% of the UKB cohort provided insufficient power to directly assess the effects of ultra-rare variants (which have <10 carriers in N~50,000 sequenced participants), we could leverage the extensive haplotype sharing within the UKB cohort^15,16^ to accurately impute these variants into up to ~100 carriers in the full cohort, thereby powering association analysis. By combining this exome-wide imputation strategy with careful fine-mapping of significant associations to identify causal effects of rare coding variants on 54 quantitative traits, we identified hundreds of novel likely-causal variant-trait associations and obtained insights into widespread allelic heterogeneity and pleiotropy.

## RESULTS

### Exome-wide imputation, association, and fine-mapping in UK Biobank

We leveraged whole-exome sequencing of 49,960 UKB participants together with SNP-array genotyping in the full cohort to impute exome-wide variants into all UKB participants as follows (full details in **Methods**). First, we created an imputation reference panel by phasing WES genotype calls together with SNP-array genotypes in the WES cohort using Eagle2^16^, restricting to 4.9 million variants with minor allele count (MAC) at least 2. Second, we used Minimac4^11^ to impute these variants into phased SNP-array haplotypes we had previously generated for 487,409 UKB participants^17^. This strategy achieved accurate imputation (*R*^2^>0.5) of rare variants down to MAF~0.00005 (**Fig. 1a, Supplementary Table 1**, and **Supplementary Note**), roughly one order of magnitude deeper into the rare allele frequency spectrum than the current UKB imputation release (v3)^1^, which used the Haplotype Reference Consortium (HRC) and UK10K / 1000 Genomes reference panels^18,19^.

**Figure 1.**
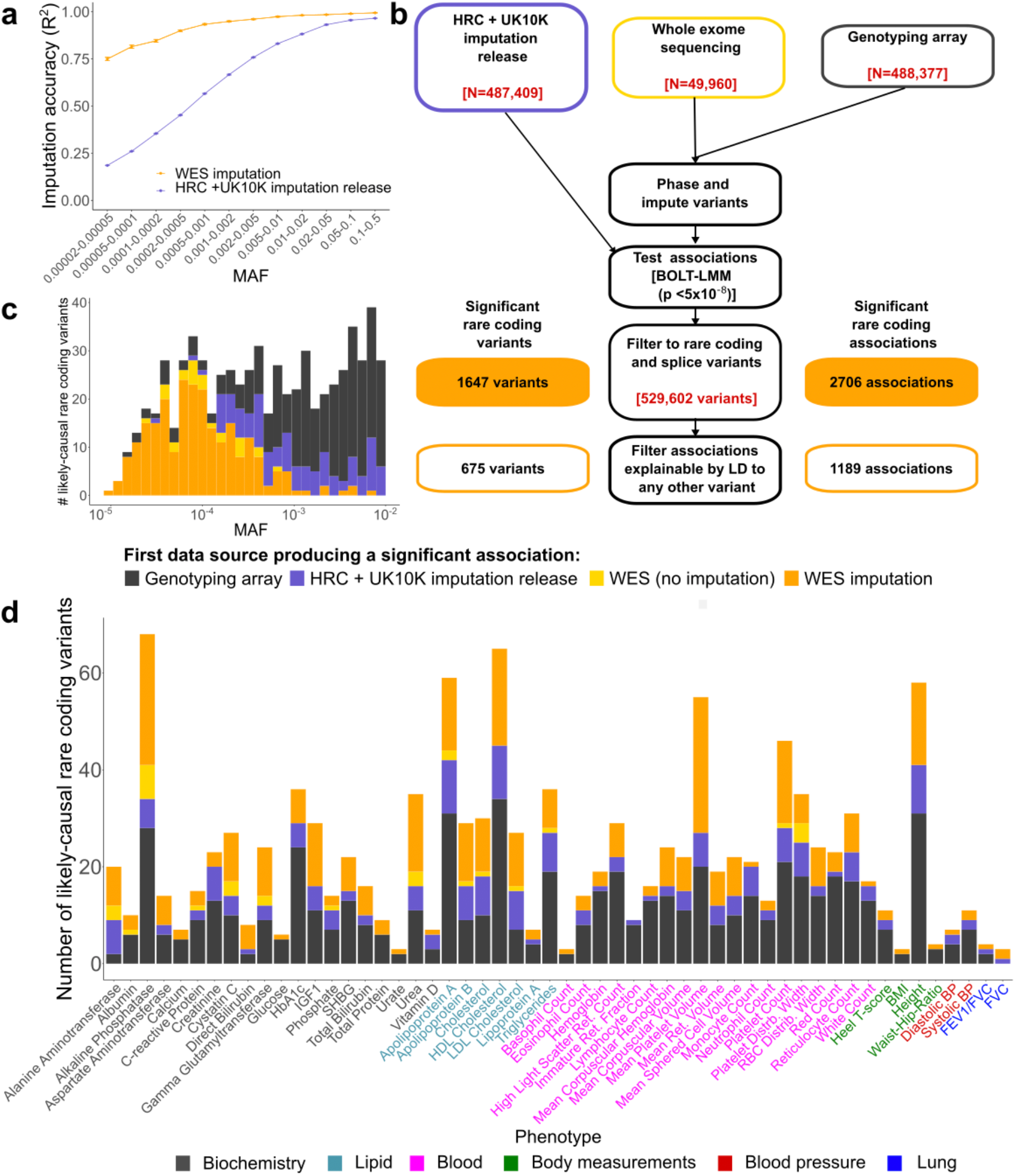
Whole-exome imputation, association, and fine-mapping identify rare coding variants likely to causally associate with 54 quantitative traits. **(a)** Imputation within UK Biobank using 49,960 exome-sequenced participants as a reference panel (orange) enables accurate imputation of much rarer variants than the HRC+UK10K-based imputation release (purple). Error bars, 95% CIs. **(b)** Schematic of our analytical pipeline, which combined UK Biobank whole-exome sequences with SNP-array genotypes to impute exome-wide genotypes into the full cohort. We analyzed imputed exome variants together with the genome-wide UK Biobank imputation release to find significant variant-trait associations independent of neighboring variants, and we restricted to rare (MAF<0.01) protein-altering variants with CADD ≥ 20 or SpliceAI support to form a final list of likely-causal variants. **(c)** Distribution of first UK Biobank genetic data set in which each association could have been detected. Roughly one-third of all likely-causal variants – and nearly all very rare likely-causal variants – were only discoverable using WES imputation. **(d)** WES imputation enabled identification of new rare coding variants for all but one trait (immature reticulocyte fraction) among 54 quantitative traits analyzed.

We tested the imputed variants for association with 54 heritable quantitative traits (measuring anthropometric traits, blood pressure, lung function, bone mineral density, blood cell indices, and serum biomarkers; **Supplementary Table 2**) by running linear mixed model association analysis on *N*=459,259 participants of European ancestry using BOLT-LMM^20,21^, which we verified was robust to potential population stratification in rare variant association analysis (**Methods** and **Supplementary Note**). This procedure identified tens of thousands of associations between coding variants and traits that reached nominal genome-wide significance (*P*<5 x 10^-8^); however, we expected that most of these associations were not causal but rather reflected linkage disequilibrium with nearby causal variants.

To filter detected associations to a high-confidence subset primarily containing causal variants, we developed a stringent filtering pipeline that combined variant annotation filters (to increase the prior on causality) with statistical fine-mapping (**Fig. 1b** and **Methods**). First, we restricted to rare (MAF<1%) variants predicted to have high protein-altering impact based on either of the following criteria: (i) Combined Annotation Dependent Depletion (CADD)^22^ score ≥ 20 (for coding variants annotated by VEP^23^, including canonical splice variants); or (ii) SpliceAI^24^ score ≥ 0.5 (for noncanonical splice variants). In our primary analyses, we further restricted to variants with high estimated imputation accuracy (INFO>0.5) and with imputed MAF>10^-5^.

These filters left 529,602 rare coding variants under consideration, of which 440,253 (83%) either were not present or were poorly imputed (INFO<0.5) in the HRC-based UKB imputation release. Among the 529,602 variants, 1,647 distinct variants associated (*P*<5 × 10^-8^) with at least one phenotype, accounting for a total of 2,706 variant-trait associations (**Fig. 1b**).

We combined our variant annotation filters with a statistical fine-mapping filter to exclude associations that could be explained by linkage disequilibrium with other variants. Our primary filter required that each association remain significant (*P* < 5 × 10^-8^, slightly conservative for 529,620 variants tested) after conditioning on any other more-strongly-associated variant within 3 megabases (considering in turn each variant from either our WES imputation or the UKB imputation v3 release; **Methods**). This filter was more robust for our rare variant analyses than standard fine-mapping software packages, which aim to find small sets of variants that explain maximal phenotypic variance, making configurations which include rare variants less likely to be considered the most probable^25,26^. Fine-mapping algorithms do have the advantage of accounting for the possibility of variants tagging combinations of multiple nearby causal variants (which our pairwise conditional filter did not consider); to account for this possibility, we applied a second filtering pipeline based on iterative runs of the FINEMAP software^25^ (**Methods**). Together, these filters reduced the set of associations to a final “likely-causal” set of 1,189 associations involving 675 unique variants (**Fig. 1b** and **Supplementary Table 3**). Both the variant annotation filters and the fine-mapping filters were designed to be very stringent, with the goal of producing a conservative set of associations with high confidence of causality for downstream analysis. Association data for all variants (including those that failed filters) are also available (see **Data availability**).

Among the 1,189 likely-causal associations, 30% could only be discovered using imputation from UKB exome-sequencing data, demonstrating the power of this approach for causal variant discovery (**Fig. 1c,d**). The remaining associations could previously have been discovered using either the UKB SNP-array (51% of likely-causal associations, reflecting the inclusion of rare coding variants on the array), the HRC-based UKB imputation v3 release (an additional 16%), or association analysis within the WES cohort (an additional 3%). Furthermore, among likely-causal associations involving ultra-rare variants (MAF<0.01%), the large majority (197 of 253 associations; 78%) were discoverable only using imputation from the UKB WES cohort (**Fig. 1c**). Most likely-causal variants (600 of 675; 89%) were not reported in the NHGRI-EBI GWAS catalog for association with any trait, underscoring the power of exome imputation within UKB to detect novel rare coding associations (**Supplementary Fig. 1**). Effect sizes generally increased with decreasing minor allele frequency among likely-causal rare coding variants (**Supplementary Fig. 2**), which collectively explained an average of 0.6% of variance per trait (**Supplementary Table 2**).

We expected that the linear mixed models we used for association tests had adequately controlled any potential confounding from population stratification or relatedness^21^. To verify robustness of our results, we performed multiple confirmatory analyses. First, we attempted to replicate associations with traits for which large-scale exome array studies (not including UKB participants) had previously been published. For height, 28 variants we identified as likely-causal had been analyzed in a previous ExomeChip study of height^2^; for all 28 variants, the direction of effect replicated, and 21 of the 28 variants reached nominal significance (*P* < 0.05) in the replication data set (**Table 1**). Similarly, effect directions replicated for 75 out of 75 lipid associations for which association statistics were available from the Global Lipids Genomics Consortium (GLGC)^3^ and for 9 out of 10 blood pressure associations for which data was available from the CHARGE-BP Consortium^4^ (**Supplementary Table 4**). Second, we verified that associations were robust to restricting analysis to a genetically homogeneous subset of unrelated British UKB participants (*N*=337,539): effect sizes (*R*^2^=0.985), association strengths (*R*^2^=0.998), and allele frequencies (*R*^2^=0.999) were all very consistent within this subset (**Methods** and **Supplementary Fig. 3**). Third, we verified that likely-causal rare alleles had geographical distributions nearly identical to MAF-matched background variants (**Supplementary Fig. 4** and **Supplementary Note**). These results indicate that while subtle stratification in large genetic analyses may affect some types of epidemiological studies^27^, the strong, highly localized stratification required to confound rare variant association analyses^28^ is unlikely to be present in UK Biobank.

**Table 1.**
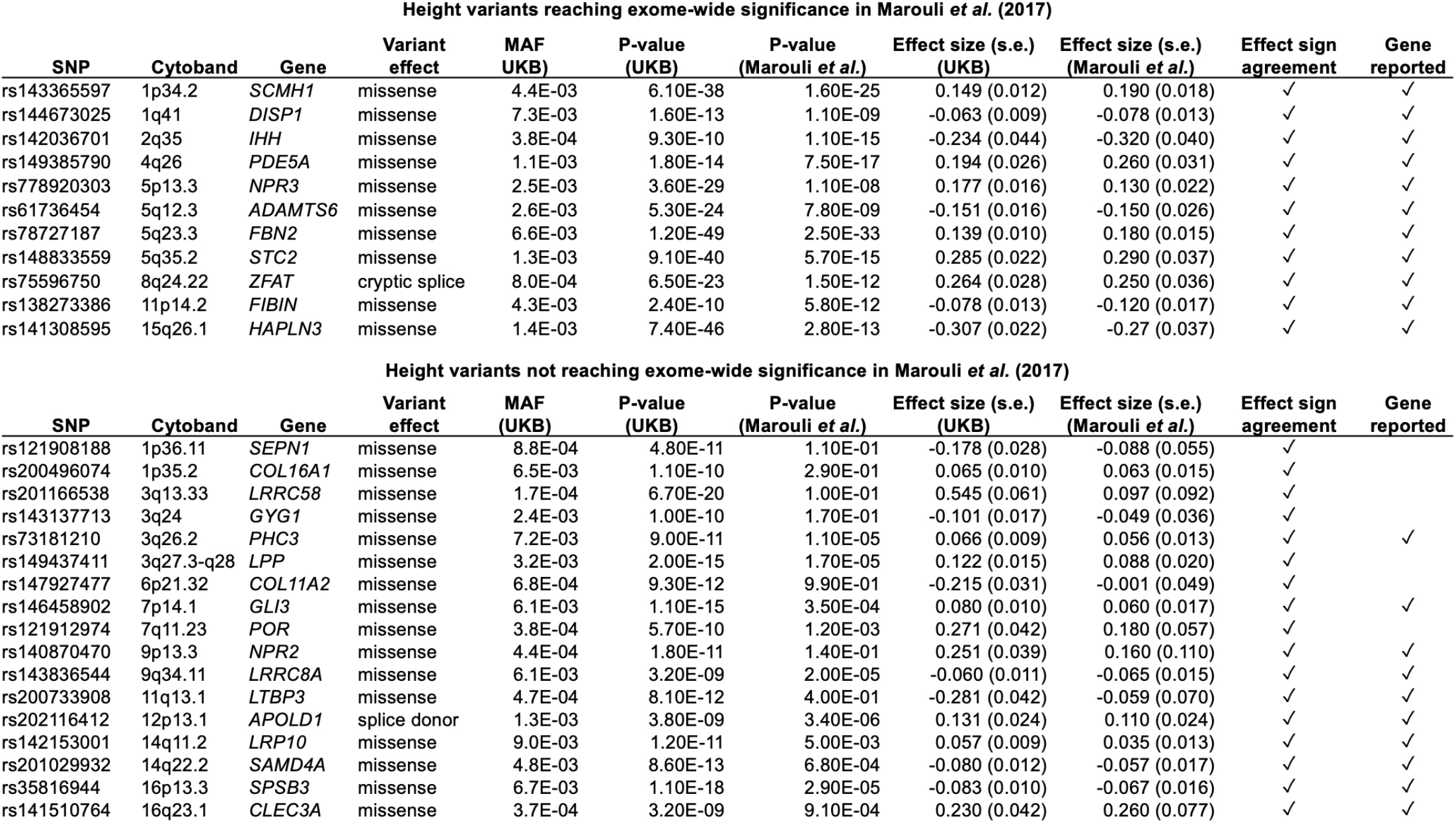
Replication of likely-causal associations between rare coding variants and height. *P*-values and effect sizes are compared for the 28 height-associated variants that were included in the ExomeChip analysis previously performed by Marouli *et al.^2^* Effect directions replicated for all 28 variants, most of which had not previously reached exome-wide significance. The last column indicates whether any variants in the affected gene had previously reached significance; several implicated genes were novel relative to Marouli *et al*.

| Height variants reaching exome-wide significance in Marouli *et al.* (2017) |  |  |  |  |  |  |  |  |  |  |
| --- | --- | --- | --- | --- | --- | --- | --- | --- | --- | --- |
| SNP | Cytoband | Gene | Variant effect | MAF (UKB) | P-value (UKB) | P-value (Marouli *et al.*) | Effect size (s.e.) (UKB) | Effect size (s.e.) (Marouli *et al.*) | Effect sign agreement | Gene reported |
| rs143365597 | 1p34.2 | SCMH1 | missense | 4.4E-03 | 6.10E-38 | 1.60E-25 | 0.149 (0.012) | 0.190 (0.018) | ✓ | ✓ |
| rs144673025 | 1q41 | DISP1 | missense | 7.3E-03 | 1.60E-13 | 1.10E-09 | -0.063 (0.009) | -0.078 (0.013) | ✓ | ✓ |
| rs142036701 | 2q35 | IHH | missense | 3.8E-04 | 9.30E-10 | 1.10E-15 | -0.234 (0.044) | -0.320 (0.040) | ✓ | ✓ |
| rs149385790 | 4q26 | PDE5A | missense | 1.1E-03 | 1.80E-14 | 7.50E-17 | 0.194 (0.026) | 0.260 (0.031) | ✓ | ✓ |
| rs778920303 | 5p13.3 | NPR3 | missense | 2.5E-03 | 3.60E-29 | 1.10E-08 | 0.177 (0.016) | 0.130 (0.022) | ✓ | ✓ |
| rs61736454 | 5q12.3 | ADAMTS6 | missense | 2.6E-03 | 5.30E-24 | 7.80E-09 | -0.151 (0.016) | -0.150 (0.026) | ✓ | ✓ |
| rs78727187 | 5q23.3 | FBN2 | missense | 6.6E-03 | 1.20E-49 | 2.50E-33 | 0.139 (0.010) | 0.180 (0.015) | ✓ | ✓ |
| rs148833559 | 5q35.2 | STC2 | missense | 1.3E-03 | 9.10E-40 | 5.70E-15 | 0.285 (0.022) | 0.290 (0.037) | ✓ | ✓ |
| rs75596750 | 8q24.22 | ZFAT | cryptic splice | 8.0E-04 | 6.50E-23 | 1.50E-12 | 0.264 (0.028) | 0.250 (0.036) | ✓ | ✓ |
| rs138273386 | 11p14.2 | FIBIN | missense | 4.3E-03 | 2.40E-10 | 5.80E-12 | -0.078 (0.013) | -0.120 (0.017) | ✓ | ✓ |
| rs141308595 | 15q26.1 | HAPLN3 | missense | 1.4E-03 | 7.40E-46 | 2.80E-13 | -0.307 (0.022) | -0.27 (0.037) | ✓ | ✓ |
| Height variants not reaching exome-wide significance in Marouli *et al.* (2017) |  |  |  |  |  |  |  |  |  |  |
| SNP | Cytoband | Gene | Variant effect | MAF (UKB) | P-value (UKB) | P-value (Marouli *et al.*) | Effect size (s.e.) (UKB) | Effect size (s.e.) (Marouli *et al.*) | Effect sign agreement | Gene reported |
| rs121908188 | 1p36.11 | SEPN1 | missense | 8.8E-04 | 4.80E-11 | 1.10E-01 | -0.178 (0.028) | -0.088 (0.055) | ✓ |  |
| rs200496074 | 1p35.2 | COL16A1 | missense | 6.5E-03 | 1.10E-10 | 2.90E-01 | 0.065 (0.010) | 0.063 (0.015) | ✓ |  |
| rs201166538 | 3q13.33 | LRRC58 | missense | 1.7E-04 | 6.70E-20 | 1.00E-01 | 0.545 (0.061) | 0.097 (0.092) | ✓ |  |
| rs143137713 | 3q24 | GYG1 | missense | 2.4E-03 | 1.00E-10 | 1.70E-01 | -0.101 (0.017) | -0.049 (0.036) | ✓ |  |
| rs73181210 | 3q26.2 | PHC3 | missense | 7.2E-03 | 9.00E-11 | 1.10E-05 | 0.066 (0.009) | 0.056 (0.013) | ✓ | ✓ |
| rs149437411 | 3q27.3-q28 | LPP | missense | 3.2E-03 | 2.00E-15 | 1.70E-05 | 0.122 (0.015) | 0.088 (0.020) | ✓ |  |
| rs147927477 | 6p21.32 | COL11A2 | missense | 6.8E-04 | 9.30E-12 | 9.90E-01 | -0.215 (0.031) | -0.001 (0.049) | ✓ |  |
| rs146458902 | 7p14.1 | GLI3 | missense | 6.1E-03 | 1.10E-15 | 3.50E-04 | 0.080 (0.010) | 0.060 (0.017) | ✓ | ✓ |
| rs121912974 | 7q11.23 | POR | missense | 3.8E-04 | 5.70E-10 | 1.20E-03 | 0.271 (0.042) | 0.180 (0.057) | ✓ |  |
| rs140870470 | 9p13.3 | NPR2 | missense | 4.4E-04 | 1.80E-11 | 1.40E-01 | 0.251 (0.039) | 0.160 (0.110) | ✓ | ✓ |
| rs143836544 | 9q34.11 | LRRC8A | missense | 6.1E-03 | 3.20E-09 | 2.00E-05 | -0.060 (0.011) | -0.065 (0.015) | ✓ | ✓ |
| rs200733908 | 11q13.1 | LTBP3 | missense | 4.7E-04 | 8.10E-12 | 4.00E-01 | -0.281 (0.042) | -0.059 (0.070) | ✓ | ✓ |
| rs202116412 | 12p13.1 | APOLD1 | splice donor | 1.3E-03 | 3.80E-09 | 3.40E-06 | 0.131 (0.024) | 0.110 (0.024) | ✓ | ✓ |
| rs142153001 | 14q11.2 | LRP10 | missense | 9.0E-03 | 1.20E-11 | 5.00E-03 | 0.057 (0.009) | 0.035 (0.013) | ✓ | ✓ |
| rs201029932 | 14q22.2 | SAMD4A | missense | 4.8E-03 | 8.60E-13 | 6.80E-04 | -0.080 (0.012) | -0.057 (0.017) | ✓ | ✓ |
| rs35816944 | 16p13.3 | SPSB3 | missense | 6.7E-03 | 1.10E-18 | 2.90E-05 | -0.083 (0.010) | -0.067 (0.016) | ✓ | ✓ |
| rs141510764 | 16q23.1 | CLEC3A | missense | 3.7E-04 | 3.20E-09 | 9.10E-04 | 0.230 (0.042) | 0.260 (0.077) | ✓ | ✓ |

### Likely-causal rare coding variants are enriched for deleteriousness

The 675 rare coding variants that we identified as likely-causal were roughly evenly distributed across the full range of allele frequencies we considered (MAF = 10^-5^ to 10^-2^; **Fig. 2a**). In contrast, the 972 rare coding variants that were annotated as high-impact and associated significantly with at least one trait but were filtered after considering linkage disequilibrium with other associated variants were enriched for more-common variants (MAF = 10^-3^ to 10^-2^), suggesting that many of these filtered variants – which constituted the majority of trait-associated rare coding variants – merely tagged causal common variants (**Fig. 2a**).

**Figure 2.**
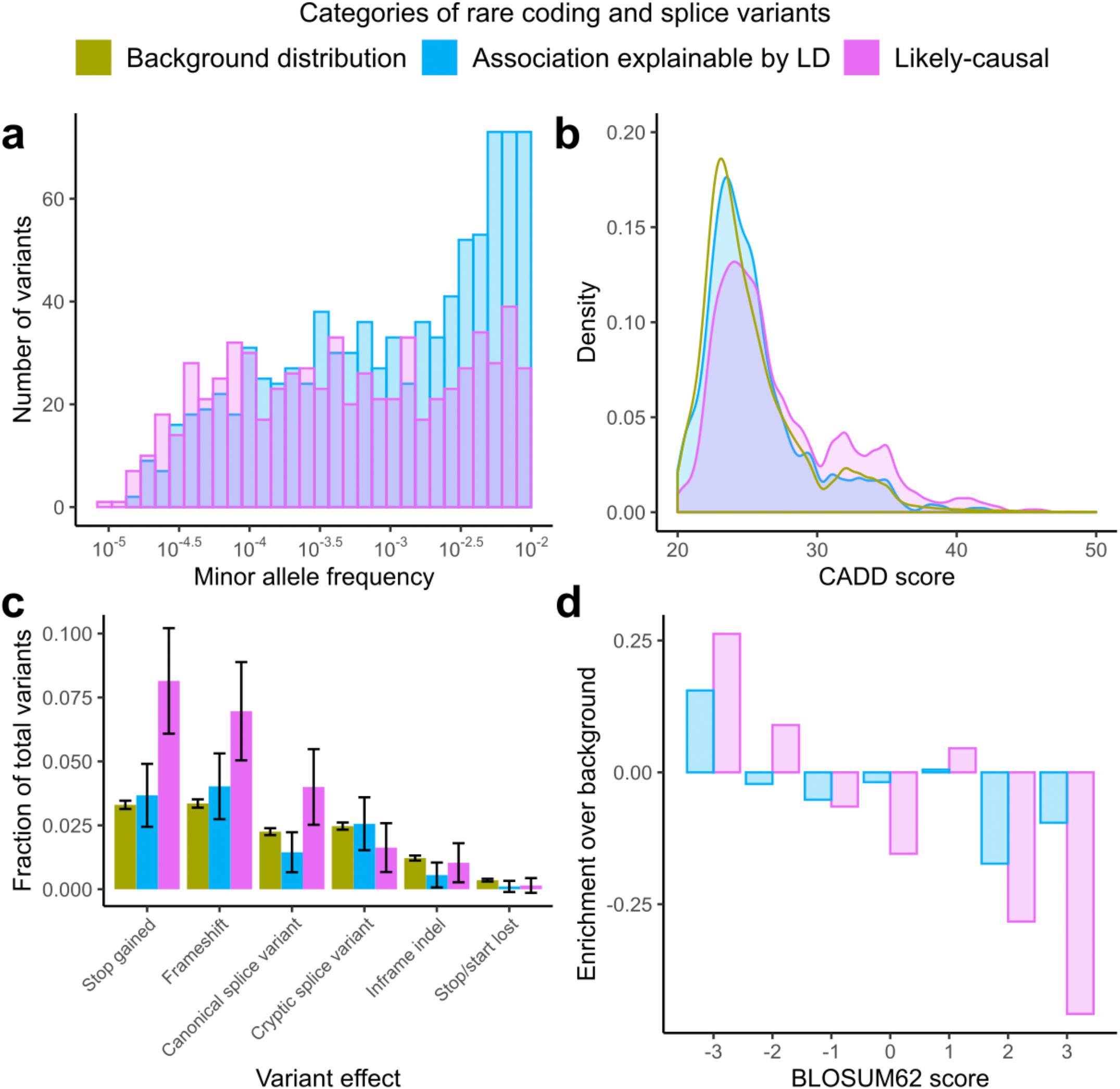
Likely-causal coding variants are rare and enriched for deleteriousness. **(a)** Likely-causal variants (pink) had minor allele frequencies distributed relatively evenly across the range under consideration (MAF = 10^-5^ to 10^-2^), whereas variants that failed linkage disequilibrium (LD)-based filters (blue) tended to be less rare. **(b)** Likely-causal variants had elevated CADD scores compared to those that failed LD-based filters and compared to a randomly-sampled background distribution of rare coding variants (green). **(c)** Likely-causal variants were enriched for predicted loss-of-function mutations. Error bars, 95% CIs. **(d)** Likely-causal missense variants were enriched for higher-impact amino acid substitutions (as measured by more negative BLOSUM62 scores).

To assess enrichment of measures of deleteriousness among the 675 likely-causal variants while controlling for MAF (which is modestly negatively correlated with deleteriousness; **Supplementary Fig. 5**), we compared features of these variants to a MAF-matched background distribution that we generated by subsampling the 529,602 predicted-high-impact variants we tested (**Methods**). The average CADD score of likely-causal variants was +1.6 higher than in the background distribution (mean CADD = 27.3 vs. 25.3; *P* = 1.6 × 10^-23^, two-sample t-test) (**Fig. 2b**). Furthermore, predicted loss of function mutations (including frameshifts, stop gains, and canonical splice variants) were 2.1-fold enriched (*P* = 3.7× 10^-16^, Fisher’s exact test) among likely-causal variants (comprising 19.1% of likely-causal variants vs. 8.9% of variants from the background distribution; **Fig. 2c**). In contrast, variants that failed our fine-mapping filters had CADD and variant type distributions similar to background, providing further evidence against causality of most of these variants (**Fig. 2b,c**). Missense variants, which comprised the majority of both likely-causal and background variants, produced broadly more severe amino acid substitutions (as measured by BLOSUM62 scores) across likely-causal variants compared to background (mean BLOSUM62 score = −0.78 vs. −0.57; *P* = 0.003, two-sample t-test) (**Fig. 2d**). Cryptic splice variants (computationally predicted by SpliceAI) accounted for 11 of the 675 likely-causal variants and were slightly depleted relative to background, suggesting that these variants were on average slightly less likely to affect function than missense variants with CADD ≥ 20 (**Fig. 2c**); however, our statistical power here was limited.

### Rare coding variants form long allelic series with consistent effect directions

Among the 1,189 likely-causal variant-trait associations we identified, roughly half (578 out of 1,189; 49%) occurred in genes containing multiple likely-causal rare coding variants for the same trait. The observation of two or more rare coding hits in the same gene strengthened our evidence for these associations and suggested the possibility of longer allelic series within these genes (containing very rare causal coding variants that either had not reached genome-wide significance or had been excluded by our stringent filters). To increase our power to detect additional independently-associated rare coding variants within these genes, we performed follow-up analyses in which we relaxed the significance threshold (to a 5% false discovery rate within each gene-trait pair) and relaxed our fine-mapping filter (conditioning only on a set of associated variants selected by FINEMAP) and annotation-based filter (considering all protein-altering variants regardless of CADD score; **Methods**).

These analyses revealed very long allelic series of rare coding variants likely to alter phenotypes: for 56 gene-trait pairs, the allelic series contained 10 or more variants on distinct haplotypes, and eight distinct genes contained allelic series of 30 or more variants (**Fig. 3** and **Supplementary Table 5**). In the longest allelic series, 45 rare coding variants in *ALPL* – out of 76 such variants tested – independently associated with serum alkaline phosphatase levels, all with negative effect directions for the rare minor allele. This consistency in effect directions was broadly displayed across the allelic series we identified (93% mean concordance with the majority effect direction; **Supplementary Fig. 6**). Somewhat surprisingly, the amino acid residues modified by missense variants within these allelic series tended not to cluster in specific protein domains (**Fig. 3a-d** and **Supplementary Fig. 7**); instead, they appeared to be distributed throughout protein structures, suggesting that protein structures may often contain many domains that are sensitive to mutation.

**Figure 3.**
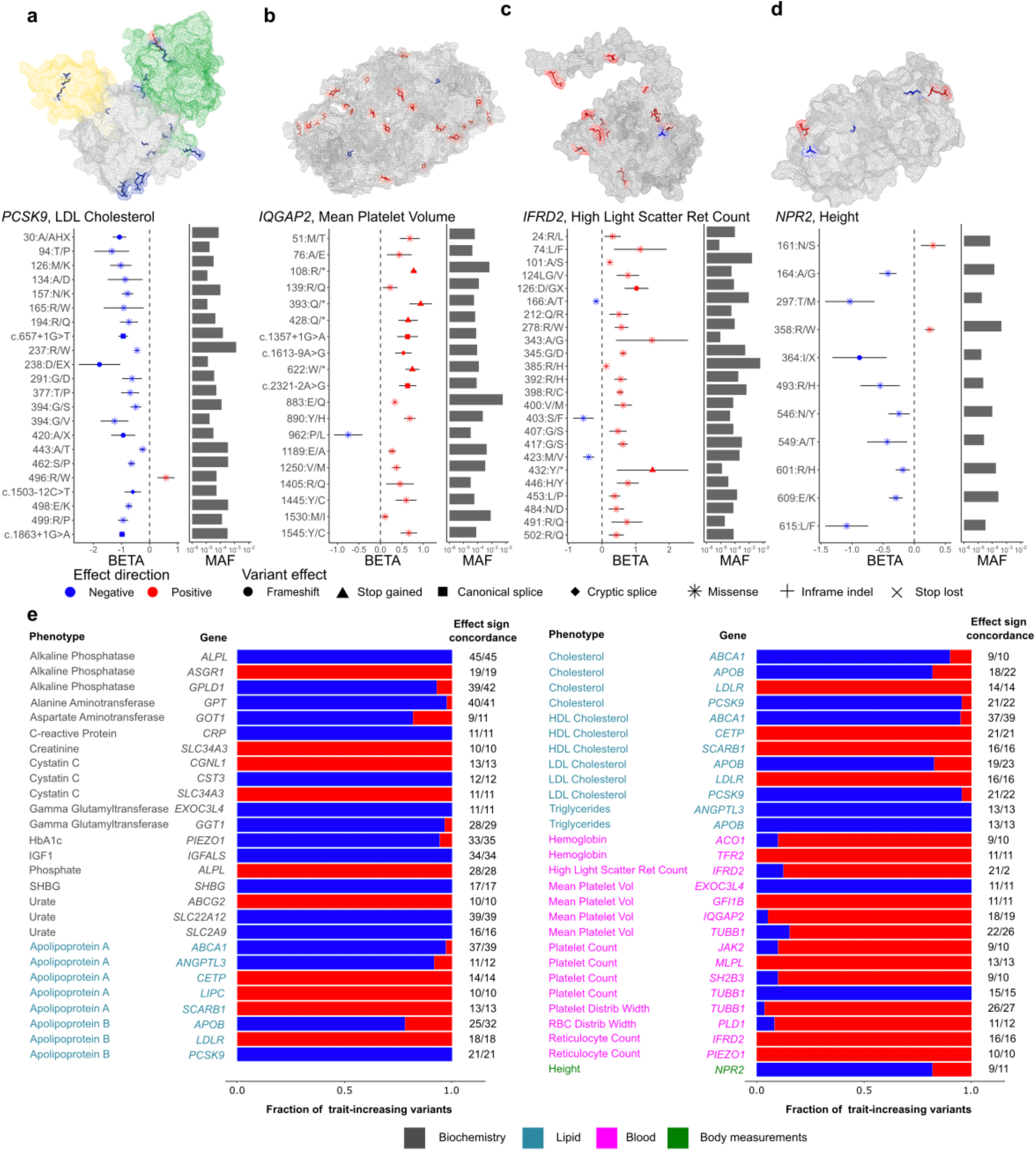
Many genes contain long allelic series of rare coding variants with consistent effect directions. (**a-d**) Allelic series of rare coding variants with statistically independent phenotype associations (reaching FDR<0.05 significance) for: (**a**) *PCSK9* and LDL cholesterol, (**b**) *IQGAP2* and mean platelet volume, (**c**) *IFRD2* and high light scatter reticulocyte count, and (**d**) *NPR2* and height. Top, protein structures with altered amino acids (modified by missense variants) color-coded by effect direction (red for trait-increasing variants and blue for trait-decreasing variants). Bottom, per-variant effect sizes (error bars, 95% CIs) and allele frequencies. Protein structures were previously determined experimentally (for PCSK9 and IQGAP2) or computationally predicted (for IFRD2 and NPR2). Functional domains of PCSK9 are shaded in different colors. IQGAP2 is represented as a homodimer in its crystal structure. (**e**) Distributions of effect directions for all gene-trait pairs with 10 or more variants in an allelic series.

Most of the allelic series we identified extended previously-described allelic series (such as in *PCSK9* and *IQGAP2*; **Fig. 3a,b**); however, several genes contained long allelic series in which most or all variants represented novel associations. At *IFRD2* (interferon-related developmental regulator 2, which has an unknown function), 24 rare coding variants independently associated with high light scatter reticulocyte count (**Fig. 3c** and **Supplementary Table 5**), suggesting an important role of *IFRD2* in red blood cell development; interestingly, these associations were specific to reticulocyte indices and did not extend to red blood cell count. A common *IFRD2* eQTL variant (rs1076872, which is synonymous in one *IFRD2* transcript and in the 5’ UTR of another transcript) exhibited the strongest association with reticulocyte indices (*P* = 1.8 x 10^-545^), and variants in linkage disequilibrium with rs1076872 have been reported by many association studies of blood cell indices. However, *IFRD2* has no common protein-altering variants, such that its apparent sensitivity to coding mutations had not previously been observable: among the 24 variants we identified, only two had MAF>0.1%. Of the remaining 22 very rare *IFRD2* variants, 19 variants had positive, large effects on high light scatter reticulocyte count (median +0.61 s.d.); intriguingly, homozygotes and compound heterozygotes for these variants exhibited extreme phenotypes (mean +2.52 s.d.; s.e.m., 0.25 s.d.).

At *NPR2*, which encodes a natriuretic peptide receptor involved in bone growth regulation^29^, 11 rare coding variants independently associated with height (**Fig. 3d** and **Supplementary Table 5**). Loss-of-function and gain-of-function variants in *NPR2* have previously been implicated in Mendelian skeletal disorders with very strong, mirror effects on stature; however, well-powered exome array studies have not linked *NPR2* polymorphisms to height in the general population^2^. Our exome-imputation approach uncovered many more *NPR2* alleles that appear to exert milder (but still strong) effects on height in the UK population, with estimated effect sizes ranging from −1.09 (0.18) s.d. to +0.25 (0.04) s.d.

At *PLA2G12A* and *PLIN1*, allelic series containing up to seven rare coding variants in *PLA2G12A* and eight in *PLIN1* associated with serum lipid levels (**Supplementary Fig. 7** and **Supplementary Table 5**), and the lead association in each series replicated in GLGC data (*PLA2G12A* missense SNP rs41278045: *P* = 3.3 × 10^-4^ for HDL and *P* = 2.3 × 10^-6^ for triglycerides; *PLIN1* missense SNP rs139271800: *P* = 1.2 × 10^-4^ for HDL). *PLA2G12A* encodes a secretory phospholipase that liberates arachidonic acid for eicosanoids with many downstream effects; *PLIN1* encodes a protein that coats lipid droplets. While frameshift variants in *PLIN1* have been implicated in Mendelian lipodystrophies^30^, the contribution of rare variants in each gene to population variation in blood lipid levels has been largely unexplored.

### Rare coding variants often exhibit pleiotropic effects

Of the 371 genes involved in at least one variant-trait association, 151 genes contained likely-causal variants for two or more traits. These associations often involved related traits or traits connected by pathways known to involve the gene in question. For example, the cell cycle regulators *CHEK2* and *JAK2* both contained likely-causal variants associated with white blood cell, red blood cell and platelet traits; a *JAK2* missense variant also associated with IGF-1 measurements (**Supplementary Table 6**). Additionally, three genes that regulate Rho GTPases *(DENND2C, DOCK8*, and *KALRN)* contained likely-causal variants associated with multiple platelet traits, consistent with the key role of Rho GTPases in platelet function^31^. Other genes associated with more-distinct sets of traits (**Supplementary Table 6**). *APOC3* exhibited the widest variety of likely-causal associations, with the splice donor variant rs138326449 associating with 13 distinct traits including lipid levels, white blood cell and red blood cell traits, and kidney biomarkers. In *PDE3B*, the stop gain variant rs150090666 associated likely-causally with 10 distinct traits, including expected associations with waist-hip-ratio and lipid measurements, but also associations with red blood cell traits, SHBG levels, and height. Further work will be required to determine which of these associations represent direct biological effects versus downstream effects of perturbed regulatory networks (as posited by the omnigenic model)^32^.

### Exome imputation uncovers novel large-effect variants for anthropometric traits

Our ability to probe the effects of ultra-rare variants revealed 10 variants in 10 different genes with very large estimated effects on height (≥ 0.5 s.d.; **Supplementary Table 7**); in contrast, the largest effect sizes detected in a recent exome array study of height were ~0.3 s.d.)^2^. Four of these genes *(NPR2, COL2A1, HERC1*, and *PCNA)* have been implicated in Mendelian diseases manifesting short stature or skeletal disorder phenotypes; however, the specific variants we identified were not previously reported in ClinVar^33^, consist with their effects being less-extreme and contributing to complex genetic variation in height. We also detected one very-large-effect variant for BMI in *MC4R* (+0.62 (0.12) s.d.; **Supplementary Table 7**); this variant had previously been associated with obesity in a Mendelian fashion^34^.

Rare coding variants with more-moderate effects on height also yielded new insights into the genetic basis of height. Among the 28 height-associated likely-causal variants for which we could replicate effect directions in the ExomeChip study of Marouli et al.^2^ (**Table 1**), seven altered genes that did not contain any variants that had previously reached significance, representing potentially novel height loci. Many of these genes had functions suggestive of their association with height, including two collagen genes, *COL16A1* and *COL11A2*. Gene Ontology (GO) analysis of all genes containing likely-causal height variants implicated numerous biological processes relating to skeletal system development and extracellular matrix organization (**Supplementary Table 8**)^35,36^.

### Biomarker-associated rare coding variants confer downstream disease risk

Many phenotypes we analyzed measured blood cell indices or biomarkers for liver, kidney, cardiovascular, or endocrine function, suggesting the possibility that rare coding variants affecting these molecular or cellular phenotypes might have downstream impacts on diseases of the corresponding systems. To test this hypothesis, we analyzed likely-causal variants from our blood and biomarker association analyses for association with disease status for related disorders (**Methods**). Seventeen associations involving 12 distinct variants reached FDR<0.05 significance (*P* < 1.5 × 10^-4^; **Supplementary Table 9**), all of which either replicated previous results^37^ or added to allelic series at known disease genes (e.g., a MAF=0.1% splice donor in *SLC34A3* that conferred threefold-increased risk of kidney stones (*P* = 2.0 × 10^-5^, OR = 3.1 (2.0-4.8)). In contrast to our analyses of quantitative traits, in which nearly one-third of the associations we identified were discoverable only through exome imputation, 11 of the 12 disease-associated variants had either been genotyped on the UKB SNP-array or accurately imputed from the HRC panel (the only exception being a MAF=0.04% *LDLR* missense variant previously implicated in familial hypercholesterolemia; **Supplementary Table 9**). This behavior was consistent with the greater difficulty of identifying robust statistical associations with disease traits (for which causal variants tend to have low penetrance) as compared to molecular or cellular traits (for which causal variants can have much more direct effects). The rarest of the 12 disease-associated variants we identified had MAF=0.04%; to identify ultra-rare variants that influence disease in population cohorts, even larger sample sizes will be needed.

### Single-variant association analyses implicate many genes missed by burden analyses

Most exome association analyses conducted to date have used gene-based association tests to aggregate signal from very rare variants within the same gene^8,9,38^, motivating a comparison between results from our single-variant analyses and a gene-based test using imputed coding variants. In light of our observation that most likely-causal variants from our single-variant analyses had consistent effect directions (**Fig. 3e**), we aggregated variants within a burden test framework (rather than using a kernel test that trades off power in this scenario to account for bidirectional effects^39^). A key consideration in performing burden tests is deciding which variants to include as potentially deleterious; as such, we considered two possible functional criteria (protein-altering with CADD≥20 vs. predicted LoF) and three possible MAF cutoffs (MAF<1%, <0.1%, or <0.01%) for variants to include (**Methods**). Of these six parameter combinations, the least stringent option (CADD≥20 and MAF<1%) appeared to be the most powerful (**Supplementary Table 10**) and was used for subsequent analyses.

Among gene-trait pairs implicated by our single-variant association tests, 32% were not detected by burden analysis, indicating that single-variant analysis can often be more powerful than gene-based tests for discovering novel loci associated with complex traits (**Supplementary Table 11**). Conversely, most gene-trait associations identified by burden analysis (1130 of 1572; 71% of associations) involved at least one variant that reached significance in single-variant analysis. Notably, a sizable minority of these associations (414 of 1130; 37% of top-associated variants) had failed our linkage disequilibrium (LD)-based filters that detected potential tagging of other causal variants, suggesting that many statistically significant results from the burden analysis could represent false-positive associations. The confounding effects of linkage disequilibrium were apparent in several large clusters of gene-trait associations near large-effect loci (e.g., 8 genes within 1 Mb of *APOE* associated with apoB levels; **Supplementary Table 11**). While burden analyses are somewhat less susceptible to confounding from LD because they aggregate signal across several variants, approximately half of the burden test associations that reached significance (51%) contained a variant accounting for the majority of alleles counted by the burden, explaining these observations. These results highlight the need to account for linkage disequilibrium even in the context of burden analysis, which may generally be better suited for case-control analyses of Mendelian traits (driven by high-penetrance, ultra-rare variants under strong negative selection) than for complex trait analyses in population cohorts.

## DISCUSSION

These results demonstrate the power of using a large, well-matched reference panel to impute very rare variants into biobank data. Whereas exome sequencing on ~50,000 UK Biobank samples offered limited power to detect associations between coding variants and phenotypes^8,9^, imputation into the remainder of the UK Biobank cohort enabled a comprehensive survey of the effects of rare coding variation on 54 quantitative phenotypes (with adequate power even for ultra-rare, MAF<0.01% variants). In combination with fine-mapping analyses, this strategy uncovered many new large-effect coding variants, revealed long allelic series within core genes for many traits, and produced a resource of likely-causal rare coding variant associations for future study. More broadly, our results suggest that sequencing 10% of a cohort and imputing into the remaining 90% can be a cost-efficient strategy for designing genetic association studies. This is true regardless of the homogeneity of the larger cohort as long as the sequenced cohort proportionally reflects the ancestral diversity present in the larger set. Accurate imputation tends to be possible for variants with at least 5-10 carriers in a reference panel^11,18^; when the panel represents 10% of a cohort, this frequency corresponds to 50-100 carriers in the full cohort, which matches well with the minimum number of carriers typically needed to detect a moderate-effect association.

Our results also have several implications for the analysis of exome association studies. First, single-variant analysis is a viable strategy for extremely large exome association studies. Second, linear mixed model association analysis is robust to population stratification for rare variants as well as for common variants. Third, careful fine-mapping is critical for identifying causal associations even when analyzing rare coding variants predicted to have high impact (CADD ≥ 20): even for such variants, most associations appear not to be causal but rather to tag associations of other variants in linkage disequilibrium (**Fig. 2**).

Our study does have important limitations. First, we restricted our primary analyses to quantitative traits; a comprehensive study of rare coding variant effects on UKB disease traits will require a separate analytical pipeline designed to handle unbalanced binary traits^40^. Second, while we could filter associations potentially explained by linkage disequilibrium with other variants imputed from exome sequencing or the HRC reference panel, we could not account for potential tagging of variants unavailable to us (e.g., very rare noncoding variants or structural variants). This limitation is shared by all fine-mapping studies conducted to date; here, we expect that our annotation-based filters (requiring that likely-causal coding variants be rare and have high predicted impact) ameliorate this concern. This intuition appears to be borne out by our replication analysis of height variants (in a pan-European meta-analysis that presumably contained different linkage disequilibrium patterns) and qualitatively by the large proportion of likely-causal associations that involved genes with clear biological relevance (**Supplementary Table 3**).

Our study of UK Biobank exome data also gives an indication of the analyses that will become feasible as exome association studies grow even larger. Very large exome-sequenced cohorts provide a natural genetic perturbation experiment: the 49,960 UK Biobank exomes we studied here contained ~7 million missense variants that modified ~3.7 million different amino acids, a sizable fraction of the ~9 million amino acids encoded by all genes in the human genome^23^. Most of these variants were singletons or doubletons and were therefore difficult or impossible to impute; however, when exome sequencing of the full UK Biobank cohort is complete, whole-exome imputation into even larger cohorts will enable characterization of the effects of much of the viable coding variation in the genome.

## METHODS

### UK Biobank genetic data

The UK Biobank cohort was previously genotyped using genome-wide SNP-arrays which produced genotype data for 488,377 UK Biobank participants at 784,256 autosomal SNPs passing quality control^12^. We analyzed these data together with whole-exome sequencing (WES) data available for 49,960 participants^8^. We analyzed WES genotype calls at 10.2 million autosomal variants from the SPB pipeline^8^, filtering to a subset of 9.8 million variants that unambiguously lifted to hg19 using UCSC liftOver, among which 4.9 million had minor allele count at least 2. We also analyzed imputed genotypes available for 487,409 participants from the UK Biobank imp_v3 data release, which consisted of 93 million variants imputed using the Haplotype Reference Consortium and UK10K / 1000 Genomes reference panels^12^.

We restricted our primary analyses to individuals who reported European ancestry (459,327 participants comprising 94% of the cohort). In supplementary analyses to ensure that our association analyses were not affected by confounding sample structure, we further restricted to a genetically homogeneous, unrelated (at third-degree or closer) subset of 337,539 white British participants^12^ (**Supplementary Note**). We excluded a small number of participants who withdrew from UK Biobank (up to a maximum of 149 withdrawals by the time we completed our study).

### UK Biobank phenotype data

We analyzed 54 heritable quantitative traits measured by UK Biobank for most participants. These traits included body measurements (3 anthropometric traits and 1 bone mineral density trait), blood pressure (2 traits), lung function (2 traits), blood cell indices (19 traits), and serum biomarker levels (7 lipid traits and 20 other biomarkers for liver, kidney, or endocrine function; **Supplementary Table 2**). We analyzed all available blood cell traits except for nucleated red blood cell count and percentage (which were mostly zero) and blood cell percentage traits (which were highly correlated with the corresponding blood cell counts). We analyzed all available serum biomarker traits except for oestradiol, testosterone, and rheumatoid factor (which had measurable levels in only half or less of the cohort). We performed basic quality control on serum biomarker traits by masking extreme outliers (>1000 times the interquartile range), stratifying by sex and menopause status, applying inverse-normal transformation, regressing out covariates (ethnic group, alcohol use, smoking status, age, height, and BMI), and re-applying inverse-normal transformation. Quality control and normalization of the other quantitative traits was previously described^21^.

We also analyzed disease traits affecting organ systems corresponding to molecular and cellular traits above. We analyzed health outcomes in the “first occurrence” data fields that UK Biobank generated by aggregating information from self-report, inpatient hospital data, primary care, or death record data.

### Phasing and imputation of WES variants

To generate an imputation reference panel from the WES cohort, we phased the 4.9 million non-singleton autosomal variants from WES together with variants genotyped on the UK Biobank array (using Eagle2^16^ with --Kpbwt=20000). We phased the data in chunks of 50,000 variants with an overlap of at least 5,000 variants between consecutive chunks, resulting in a total of 126 chunks across all autosomes. We then imputed the WES-derived variants into phased haplotypes we had previously generated^17^ for 487,409 participants in the full cohort (using Minimac4^11^ with noncoding variants from the UK Biobank array used as the imputation scaffold). We benchmarked the accuracy of this imputation approach by computing correlations between imputed genotype dosages and direct genotype calls at well-typed coding variants included on the UK Biobank genotyping array, which were excluded from the imputation scaffold (**Supplementary Note**).

### Association tests

We tested variants for association with each of the 54 quantitative traits using the non-infinitesimal linear mixed model association test implemented in BOLT-LMM^20^ (--lmmforceNonInf) with assessment center, genotyping array, sex, age, age squared, and 20 genetic principal components included as covariates. We fit the mixed model on directly-genotyped autosomal variants with MAF>10^-4^ and missingness<0.1 and computed association test statistics for WES-imputed variants and variants from the UK Biobank imp_v3 release. In our primary analyses, we included all participants with non-missing phenotypes who reported European ancestry (and had not withdrawn from the study). We also performed association analyses that further restricted the sample set to the WES cohort to determine which associations were detectable in the WES cohort alone.

### Filtering associations using coding variant annotations

To focus our analyses on variants likely to have protein-altering effects, we filtered significant associations to those involving variants predicted (by genome annotation algorithms) to impact function. For variants modifying protein-coding sequence or canonical splice sites, we required a CADD v1.3 score ≥ 20 and a VEP annotation of missense, inframe deletion, inframe insertion, start lost, stop lost, splice acceptor, splice donor, frameshift, or stop gained^22,23^. For variants that affected multiple transcripts (for one or more genes), we assigned the most severe VEP annotation (in the order listed above) across all affected transcripts. We also included potential cryptic splice variants predicted by SpliceAI v1.2 (specifically, variants with a delta score ≥ 0.5 for at least one of the four splice modifier categories: gain or loss of a splice acceptor or a splice donor)^24^.

### Filtering associations potentially explained by linkage disequilibrium with more-strongly-associated variants

To further filter significant associations to a high-confidence set of likely-causal associations, we analyzed linkage disequilibrium (LD) between pairs of associated variants to identify and remove any associations potentially attributable to tagging of another variant in LD. We took this approach because while many algorithms have been developed for fine-mapping common variant associations, these methods are not optimized for rare variants: intuitively, they maximize the heritable variance that can be explained by a configuration of causal variants, making configurations which include rare variants – which typically account for very little heritability even though they can have large effect sizes – less likely to be considered probable^25,26^.

Our filter, which was equivalent to requiring that each association remain significant (*P* < 5 × 10^-8^) after conditioning on any other more-strongly-associated variant nearby, proceeded as follows. For each rare coding variant *i* significantly associated with a phenotype, we calculated its correlation *r_ij_*(i.e., in-sample LD) with each other more-strongly-associated variant *j* (including both WES-imputed variants and variants from the HRC-based imputation release) using plink “--r”^41^. We then computed the approximate chi-square statistic that would be obtained for variant *i* in a model including variant *j* as a covariate:

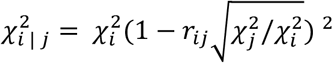

where 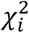 and 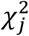 denote the chi-square test statistics computed by BOLT-LMM for variants *i* and *j*. In order to retain variant *i*’s association as likely-causal, we required the conditional chi-square statistic 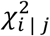 to exceed 29.7168 (corresponding to *P* < 5 × 10^-8^) for every variant *j* with 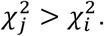

Filtering associations potentially explained by linkage disequilibrium with multiple variants.

The filter described above was designed to eliminate associations involving variants that primarily tagged one other variant in LD; however, in theory, non-causal variants could escape this filter by tagging a combination of multiple other variants. To account for this possibility, we used the FINEMAP software^25^ to determine, for each gene harboring a rare coding variant of interest, whether the local genetic architecture appeared to involve multiple causal variants, and if so, to assess whether the rare coding variant(s) under consideration remained significantly associated after conditioning on the variants selected by FINEMAP.

We performed this analysis using a two-step procedure. First, we ran FINEMAP’s shotgun stochastic search algorithm (“--sss”) to identify up to 5 putatively causal variants among all significantly associated variants within 500kb of the gene under consideration. This run produced a most probable configuration of 1-5 variants, most of which were typically common. We then ran FINEMAP a second time, adjusting the number of allowed causal variants to be one greater than the number selected for the top configuration in the first run, and limiting the set of potential causal variants to those variants in the top configuration from the first run along with all significantly-associated rare coding variants in the gene under consideration. The purpose of this second run was to ascertain whether each rare coding variant remained significant in a model conditioning on multiple common variants. Specifically, we extracted the conditional z-scores output by FINEMAP in its “.snp” files and dropped variants with z-score ≤ 4. This filter only removed 20 variants involved in 36 associations, suggesting that most rare variants that tagged other causal variants were primarily tagging just one neighboring variant. We set the z-score threshold to ≤ 4 after exploring other cut-offs such as *z* ≤ 5.45, the equivalent of a genome-wide significance threshold. The *z* ≤ 5.45 threshold filtered an additional 54 variants; however, several associations with z-scores around 5 that failed this filter appeared to be real (e.g., high-CADD or stop gain mutations in genes known to alter lipid levels). In light of this observation and the stringent filtering we had already performed using pairwise tests, we decided to set a threshold of *z*-score ≤ 4, which appeared to filter primarily spurious associations. Applying this filter together with the previous two filters left us with the final list of 1,189 significant rare coding variant associations involving 675 unique variants for 54 quantitative traits.

### Variant lookup in the NHGRI-EBI GWAS Catalog

We compared the variants we identified to those reported in the NHGRI-EBI GWAS Catalog (accessed January 15, 2020)^42^. Each variant was checked to see if it was reported in the catalog for any phenotype to exclude the possibility that the variant was previously reported for a related phenotype.

### Replication analyses

Several traits we analyzed had previously been studied in large-scale meta-analyses using exome arrays, providing the opportunity for replication of likely-causal associations that involved variants assayed on the exome arrays. We compared the associations of likely-causal variants we identified for height, blood pressure, and lipid measurements (LDL cholesterol, HDL cholesterol, triglycerides, and total cholesterol) to association statistics previously published by the GIANT Consortium^2^ (N=381,625), the CHARGE-BP Consortium^4^ (*N*=120,473), and the Global Lipids Genetics Consortium (*N*≈300,000), respectively^3^; all of these meta-analyses predominantly studied participants of European ancestry, and none included UK Biobank. While most variants were too rare to attain statistical significance in these replication data sets (probably due to allele frequency differences between the UK and other European populations), 112 out of 113 associations exhibited the same effect direction in UK Biobank and the replication data set (**Table 1** and **Supplementary Table 4**). We also compared our height associations to association statistics reported from exome-sequencing of the FinMetSeq cohort^43^ (*N*=19,241), which provided replication support for a few additional variants that happened to have higher allele frequencies in Finns (**Supplementary Table 4**).

### MAF-matched background distribution for assessing deleteriousness enrichment

To identify trends in the deleteriousness of likely-causal rare coding variants as compared to all rare coding variants, we generated a background distribution of rare coding variants with a MAF distribution matching that of the likely-causal variants (to account for the tendency of rarer variants to have higher deleteriousness scores). We first stratified likely-causal variants into three MAF bins: 10^-5^-10^-4^, 10^-4^-10^-3^, and 10^-3^-10^-2^. We then subsampled the set of all rare coding variants considered in our analyses (regardless of whether or not they had a significant association) using the R “sample” function to generate a set of variants with the same fraction of variants in each MAF bin as in the likely-causal set. We included all variants in the highest MAF bin (as this bin contained the fewest variants), which set the total number of variants in the background distribution at 47,142 variants.

### Allelic series analyses

As our primary analysis pipeline for identifying likely-causal rare coding variant associations implemented strict filters on statistical significance (in both single-variant analysis and conditional analyses), we applied a secondary analysis pipeline that relaxed these filters to identify additional rare coding variant associations with good statistical support within genes with two or more likely-causal variants for a trait (indicating strong evidence for the gene-trait association). This pipeline applied a two-step approach (detailed in the **Supplementary Note**) using FINEMAP in a manner somewhat similar to the approach we used to filter associations that could be explained by combinations of other variants. Here, we again performed a first run of FINEMAP to allow it to select a multiple-causal-variant model (this time containing up to 15 causal variants chosen from common and low-frequency variants as well as rare coding variants), and we then ran FINEMAP a second time to perform an iterative conditional analysis using the selected variants together with rare coding variants. We used conditional P-values from the second FINEMAP run to assess the extent to which each rare coding variant exhibited a trait association independent of previous variants. Finally, we converted P-values to g-values to determine the set of rare coding variants that reached significance at a false discovery rate of 5%.

The expanded allelic series we identified at FDR<0.05 significance often contained many variants. (For genes with multiple transcripts, we counted the lengths of the allelic series for the transcript that contained the most FDR<0.05-significant variants, treating cryptic splice variants as belonging to all transcripts.) To visualize the effects of missense variants, we plotted the affected amino acids on previously-generated protein structures. Experimentally-derived protein structures for PCSK9 (2P4E), ANGPTL3 (6EUA), IQGAP2 (5CGJ), and GOT1 (3II0) were retrieved from PDB^44^. Computationally predicted structures for NPR2 (P20594 monomer) and IFRD2 (Q12894 monomer) were retrieved from SWISS-MODEL^45^.

### Associations with health outcomes

We tested likely-causal variants we identified for cellular and molecular phenotypes (blood cell traits, liver biomarkers, diabetes biomarkers, renal biomarkers, and cardiovascular biomarkers) for associations with corresponding disease outcomes coded by UK Biobank using ICD-10 codes (blood disorders, D50-D77; liver diseases, K70-K77; type 2 diabetes, E11; gout and kidney diseases, M10 and N00-N29; cardiovascular diseases, I20-I25 and I63). To further reduce multiple testing burden, we further restricted to diseases with at least 500 reported cases. These criteria left 40 phenotypes under consideration (i.e., an average of 8 phenotypes tested for each likely-causal variant for each of the 5 classes of cellular/molecular phenotypes) and resulted in 5,508 separate tests. Setting a false discovery rate threshold of 5% across the 5,508 tests resulted in a significance threshold of *P* < 1.5 × 10^-4^.

### Gene-based burden tests

We assessed the performance of gene-based association analyses using burden tests that collapsed the genotypes of imputed rare coding variants within each gene. We considered six different criteria for inclusion of rare coding variants in the burden. These six criteria were defined by three different allele frequency thresholds (MAF ≤ 1%, 0.1%, and 0.01%) and two different variant annotation criteria (protein-altering with CADD ≥ 20 or predicted loss-of-function as annotated by VEP). Collapsed genotypes were coded as 0 (if an individual had no variants meeting these requirements) or 1 (if the individual carried at least one of these variants). We performed association tests against the 54 quantitative traits using BOLT-LMM with the same settings as in our single-variant analyses, and we applied a Bonferroni-corrected *P*-value threshold of *P* < 2.7 × 10^-6^ to account for 18,530 genes tested. We compared the results of these analyses to those previously reported in burden analyses of *N*=49,960 exome-sequenced UK Biobank participants^8,9^. Among phenotypes in common between our analyses and the previous analyses, we replicated 13/15 associations from Van Hout *et al*.^8^ and 48/58 associations from Cirulli *et al*.^9^. Non-replicated results might arise from different selection criteria for variants and to a lesser extent from singletons that were included in the previous analyses but excluded from our imputation.

### Data availability

Access to the UK Biobank Resource is available by application (http://www.ukbiobank.ac.uk/). Exome-wide summary association statistics for the 54 quantitative traits we analyzed are available at https://data.broadinstitute.org/lohlab/UKB_exomeWAS/, and data files containing allelic series for all gene-trait associations with multiple likely-causal variants are also available at this website.

### Code availability

The following publicly available software packages were used to perform analyses: Eagle2 (v2.3.5), https://data.broadinstitute.org/alkesgroup/Eagle/; Minimac4 (v1.0.1), https://genome.sph.umich.edu/wiki/Minimac4; BOLT-LMM (v2.3.4), https://data.broadinstitute.org/alkesgroup/BOLT-LMM/; FINEMAP (v1.3.1), http://www.christianbenner.com/; plink (v1.9 and v2.0), https://www.cog-genomics.org/plink2/. Scripts used to perform the downstream analyses described above are available from the authors upon request.

## Data Availability

Access to the UK Biobank Resource is available by application (http://www.ukbiobank.ac.uk/). Exome-wide summary association statistics for the 54 quantitative traits we analyzed are available at
https://data.broadinstitute.org/lohlab/UKB_exomeWAS/, and data files containing allelic series for all gene-trait associations with multiple likely-causal variants are also available at this website.

http://www.ukbiobank.ac.uk/

## Acknowledgments

We thank A. Gusev, M. Hujoel, P. Palamara, A. Price, and S. Sunyaev for helpful discussions. This research was conducted using the UK Biobank Resource under application #10438. A.R.B. was supported by training grant T32 HG 2295-16. M.A.S. was supported by the MIT John W. Jarve (1978) Seed Fund for Science Innovation. R.E.M. was supported by US NIH grant K25 HG150334. P.-R.L. was supported by US NIH grant DP2 ES030554, a Burroughs Wellcome Fund Career Award at the Scientific Interfaces, the Next Generation Fund at the Broad Institute of MIT and Harvard, and a Sloan Research Fellowship. Computational analyses were performed on the O2 High Performance Compute Cluster, supported by the Research Computing Group, at Harvard Medical School (http://rc.hms.harvard.edu).

## Author contributions

A.R.B and P.-R.L. performed statistical analyses and wrote the manuscript. M.A.S. and R.E.M. provided substantial input on all analyses and on the manuscript.

